# Use of Environmental Variables to Predict SARS-CoV-2 Spread in the U.S.

**DOI:** 10.1101/2021.05.19.21257350

**Authors:** R. Sterling Haring, Sean Trende, Christina M. Ramirez

**Affiliations:** Department of Physical Medicine and Rehabilitation, Vanderbilt University Medical Center, Nashville, TN; Department of Health Policy and Management, Johns Hopkins Bloomberg School of Public Health, Baltimore, MD; Department of Political Science, Ohio State University, Ohio, United States of America; Department of Biostatistics, Jonathan and Karen Fielding School of Public Health, University of California, Los Angeles, California, United States of America

**Keywords:** COVID-19, viral spread, modeling, temperature, Bayesian heirarchical models

## Abstract

**Background:** The COVID-19 pandemic has challenged even the most robust public health systems world-wide, leaving state and local health departments, hospitals, and physicians with little guidance on planning and resource allocation. Efforts at predicting the virus’ spread have largely failed to capture the nuances presented by national and local geographic, environmental, and sociological variables.

**Objective:** Using county-level data from the United States, we sought to measure the extent to which these demographic, geographic, and environmental variables correlate with the spread of COVID-19.

**Methods:** Using demographic data from the US Census Bureau’s American Community Survey, weather station data from the National Oceanic and Atmospheric Administration (NOAA), and COVID-19 case data from the Center for Systems Science and Engineering at Johns Hopkins University and the New York State Department of Health, we employed Bayesian hierarchical modeling with zero-inflated Negative Binomial regression to calculate correlations between these variables, COVID-19 case count, and rate of viral spread. Key predictors were identified and measured during two periods of two weeks each: March and June of 2020. The resultant model was then employed to predict case counts and spread rate for early July 2020.

**Results:** While demographic and environmental factors explain viral spread well, our findings challenge earlier conclusions about how these factors related to viral progress. Using these factors alone, we were able to predict spread to within 1% in all but 8 counties (99.9%), and within 0.1% in all but 51 counties (98.4%). The model was subsequently able to predict early July viral spread to within 0.5% in 98% of counties. Contrary to earlier findings, temperature had variable effect; as Spring temperatures warmed, cases decreased, but Summer heat increased cases, likely reflecting movement of populations from indoors to outdoors and back in. States varied little in their case rate relative to the model, and much of the variation could be linked to known “superspreader” events.

**Conclusion:** While environmental and demographic variables can help predict COVID-19 spread rates, some relationships are variable in ways earlier research failed to identify.

**Role of Funding Source:** There was no funding supporting this work.

## 1. Introduction

COVID-19 has infected over 81 million people and killed over 1,700,000[1]. Due to the rapid spread of the novel virus, governments and physicians have had to manage the clinical, societal, and economic ramifications with imperfect data. While certain aspects of the virus are well-understood, little is known about the macro-environmental factors contributing to viral spread.

Early in the pandemic, researchers believed that warm weather could slow the spread of the virus [2]. This was buttressed by a few early studies suggesting that warm weather and high humidity slowed spread. Wang et al. examined the spread of COVID-19 in 100 Chinese cities and 1,005 U.S. counties, and found that temperature and relative humidity had a significant, negative effect on viral transmission[3]. They concluded that viral spread would likely slow in the summertime, although temperature alone would not be enough to reduce the R0 below one. Other work reached similar conclusions[4, 5].

In the US, outbreaks were, in fact, limited in northern states for much of the summer. States like Arizona, Florida and Texas, however, saw spikes of the virus, seemingly confounding earlier expectations[6]. In addition, countries with hot summer climates such as Brazil, Mexico, and Peru saw the disease spread widely[7]. This summer wave has attracted less analysis that the first wave, despite the tension it creates with theories of viral spread in the earlier literature.

To help better understand the relationship between COVID spread and environmental and demographic factors, we examine COVID-19 spread in the US during what we see as two distinct phases: the initial outbreak of the virus (mid-March through early April), and the second (mid-June through early July).

## 2. Methods

The two time periods of interest were selected based on a combination of factors. We chose the first time period, measured with what we call the “Spring Model,” because it represented a time where COVID-19 had spread widely enough to create a reasonable amount of variance in the data, and because infections that presented during this time period were likely transmitted before or shortly after U.S. President Donald Trump declared a state of emergency[8] and as states were implementing a variety of non-pharmaceutical interventions [9, 10]. In other words, we selected a time period when viral spread in all states was accelerating.

We also modeled spread in late June with our “Summer Model.” We selected this time period because most states had begun to emerge from lockdowns by this point, reducing between-state variation[11]. As a sensitivity and robustness analysis, we used the Summer model to try and predict viral spread completely out-of-sample, during the month of July.

### 2.1. Data

We took daily case counts from the Center for Systems Science and Engineering (CSSE) at Johns Hopkins University[12]. CSSE data do not break New York City down by borough, so we took data from the New York State Department of Health COVID-19 Tracker[13]. We obtained county-level population estimates from the US Census Bureau’s American Community Survey (ACS) for 2019 [14], and calculated population density by dividing these estimates by the land area of each county as reported by the US Census Bureau[15]. Data collected from cruise ships and US territories were excluded.

Humidity data were taken using weather station data from the National Oceanic and Atmospheric Administration (NOAA) [16]. We identified daily temperature and dew point data for each weather station for the months of interest and calculated the mean humidity for each month using the Magnus formula [17]. We then calculated the distance from each county centroid to each weather station, and data from the nearest weather station were assigned accordingly.

County-level temperature data were not available on a day-to-day basis, and including such data in the model would have been difficult given the lag between infection and diagnosis[18]. Instead, we use the mean county temperature for the months of interest, under the assumption that post-transmission lag in presentation would be relatively normally distributed, and that cases transmitted in early- and mid-month would present around mid- and late-month, respectively [19].

We extracted latitude and longitude for the centroids of each county from CCSE data, and included these as covariates. Latitude was included to test hypotheses about the effects of sunlight on viral transmission. Longitude was included to control for the observation that the early outbreaks of the virus tended to occur on the U.S. coasts, and then spread inward[20]. To capture and quantify this phenomenon, we calculated the absolute distance of the longitude for the centroid of each county from the central meridian of the 48 contiguous states (−98.583 degrees).

Our primary outcome was the change in cases in all 3,108 United States counties located within the contiguous 48 states from March 16, 2020 to April 1, 2020 (Spring model), and from June 16, 2020 to July 1, 20020 (Summer model). A handful of counties showed a decrease in cumulative cases over these three weeks; these were dropped, as they almost certainly reflect reporting errors or estimate revisions.

Because we utilize a difference-in-differences estimator for our response, we were sparing in our control variables. Because New York City was hit hard early in the pandemic, separated Metropolitan New York City into two variables: one including only the five-county New York City area, and another excluding this area but including the rest of the area making up the Metropolitan New York City Statistical Area. Finally, we controlled for relative humidity, following the approach of Wang et al. [3]

Because data are publicly available and no personally identifying characteristics are available in the data set, this study was exempt from Institutional Review Board approval.

### 2.2. Statistical Analysis

We estimated factors driving the rate of viral spread utilizing a Bayesian hierarchical model, estimated through the R-INLA package[21], using nested Laplace approximations to obtain fast calculations of Bayesian hierarchical models[22]. Case counts are necessarily positive (since cumulative infection numbers cannot decrease over time), so we utilized a generalized linear model with a logarithmic link[23].

To test for possible non-linearities in the data, population densities were squared and cubed.

Zeroes dominate the distribution of case spread, accounting for about 1/3 of the data, possibly due to limited testing or undetected cases (Figure 1). To account for this, we employed a zero-inflated model, which estimates an additional parameter to account for the possibility that a zero represents a “false negative.” To better conceptualize this approach, one might imagine that the number of fish caught by people fishing in a park follows a Poisson distribution. If one estimates a simple Poisson model by counting the number of fish caught by everyone exiting the park, you may find an excessive number of zeroes because some people exiting won’t have fished at all. The zero number of fish caught, in this sense, is false. In other words, there is actually a two step process involved: a process for determining whether someone opts to fish, and a process for estimating the number of fish caught by those who do fish.

**Figure 1:**
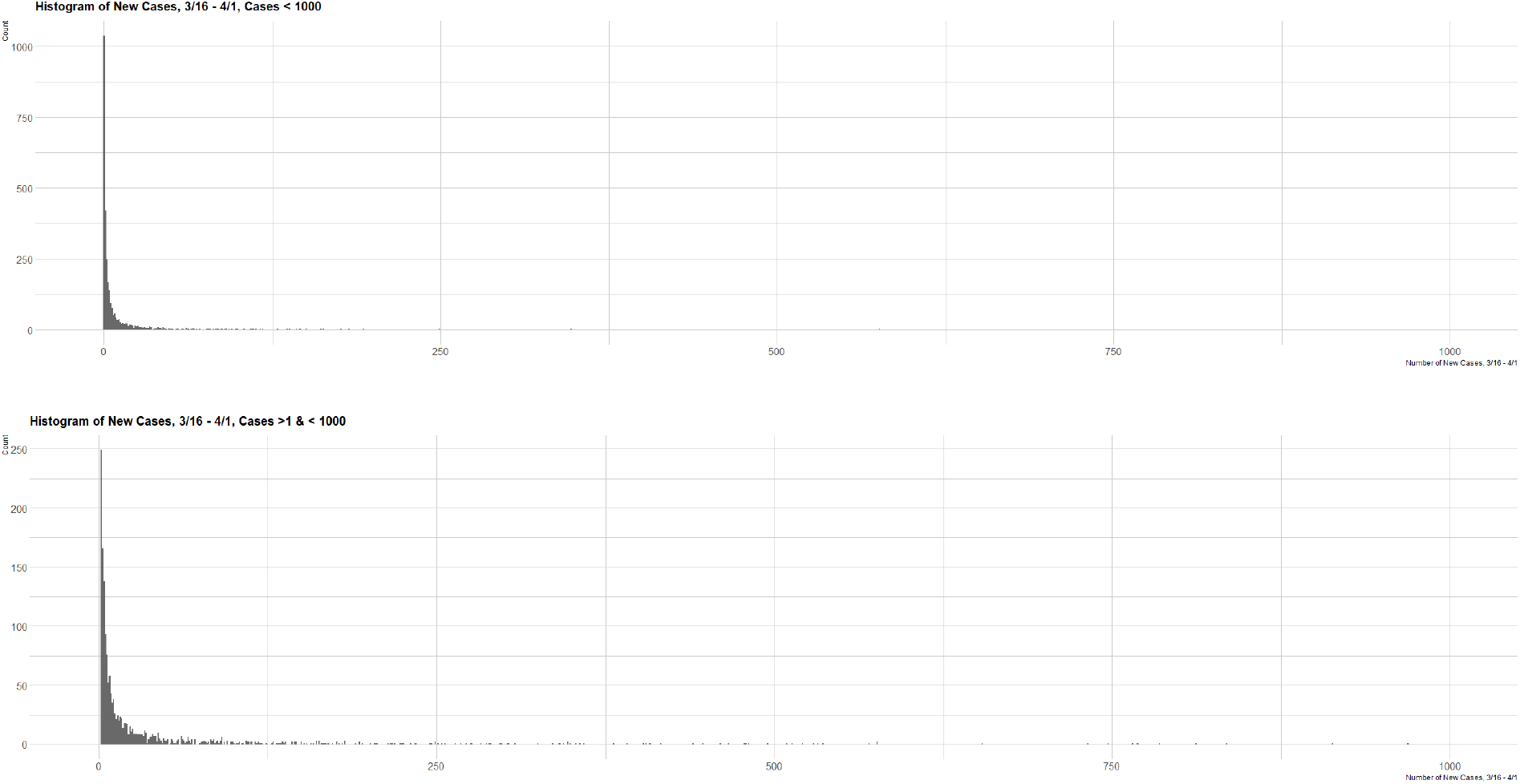
Distribution of Response Variable, With and Without Zeroes

The zero-inflated model therefore first estimates the probability that a given zero is false. It then estimates the remaining terms using a different distribution. Due to overdispersion, we used a (zero-inflated) negative binomial regression model.

All calculations were performed using the R Statistical Package, version 3.6.2 [24].

## 3. Results

### 3.1. Spring model

In the Spring wave, the New York Metro area showed a strong correlation with case spread (1.348, 95% CI: 0.805, 1.932), suggesting that, even after applying controls, the New York Metro area suffered from an unusually high rate of infection in early Spring. Longitudinal distance from the central meridian of the United States also showed a positive correlation (0.057, 95% CI: 0.038, 0.078), consistent with viral spread inward from the coasts. Latitude was inversely correlated (−0.11, 95% CI: -0.162, -0.059), suggesting that counties that were further North had a lower rate of case increase than counties further South, after controlling for other variables.

Relative humidity was not a significant predictor of viral spread (0.00457, 95% CI: -0.002, 0.001), contradicting findings from Wang, et al. Mean temperature in the Spring was inversely correlated with case spread (−0.025, 95% CI: -0.044, -0.006), suggesting that states did benefit from warm weather during this time, after adjusting for other factors.

Population density showed an exponential positive effect on spread (4.85e-04, 95% CI: 3.927e-04, 5.842e-04).

Rates of viral spread not accounted for in our model, which may include local and regional factors such as shutdown orders and the availability of testing in a given area, are illustrated in Fig. 2. Counties located in Colorado, Idaho, Louisiana, North Dakota and Washington seemed to have unusually high rates of Spring COVID-19 spread, even after controlling for other factors, while California, Florida, Kentucky, Maine, North Carolina, Virginia and West Virginia seem to have unusually low levels of spread during the Spring. Overall, half of all states had no significant deviation from case rates predicted by the Spring model. As suggested by the high accuracy of prediction at the state level, overall fit for the Spring model was quite good. Examining the residuals, half of all counties were predicted within a single case, 83% of counties were predicted within 10 cases, and 97.2% of counties were predicted within 100 cases. Of course, there is a substantial difference between predicting within 100 cases in a county of 1,000 people, and predicting within 100 cases in a county of 1,000,000. To account for this, we examined rate of spread by dividing the residuals by the population, and examine the rate of spread directly. The Spring model predicted the rate of spread in counties well: within a percentage point in all but 8 counties (99.9%) and within a tenth of a point in all but 51 counties (98.4%).

**Figure 2:**
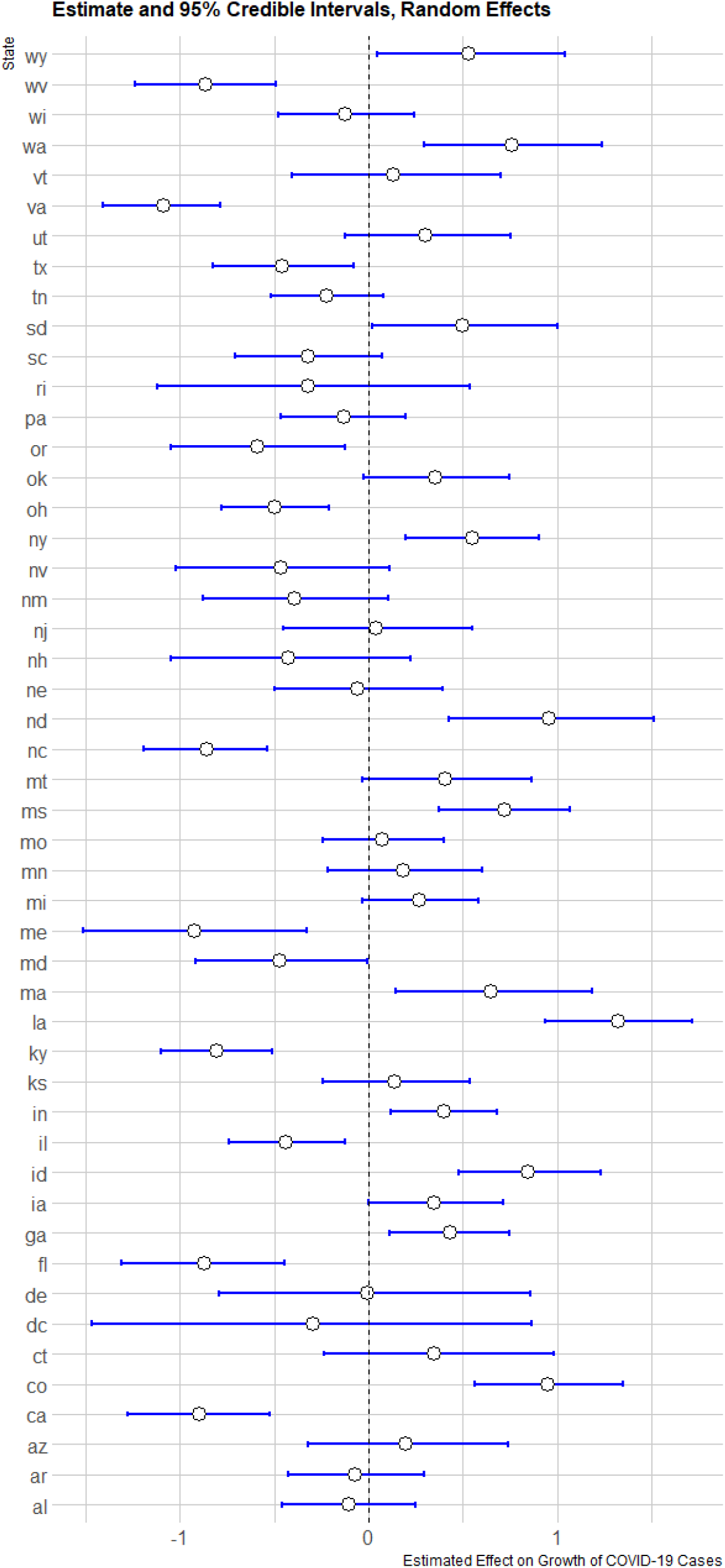
Random Effects for COVID Spread in States, late March 2020

**Figure 3:**
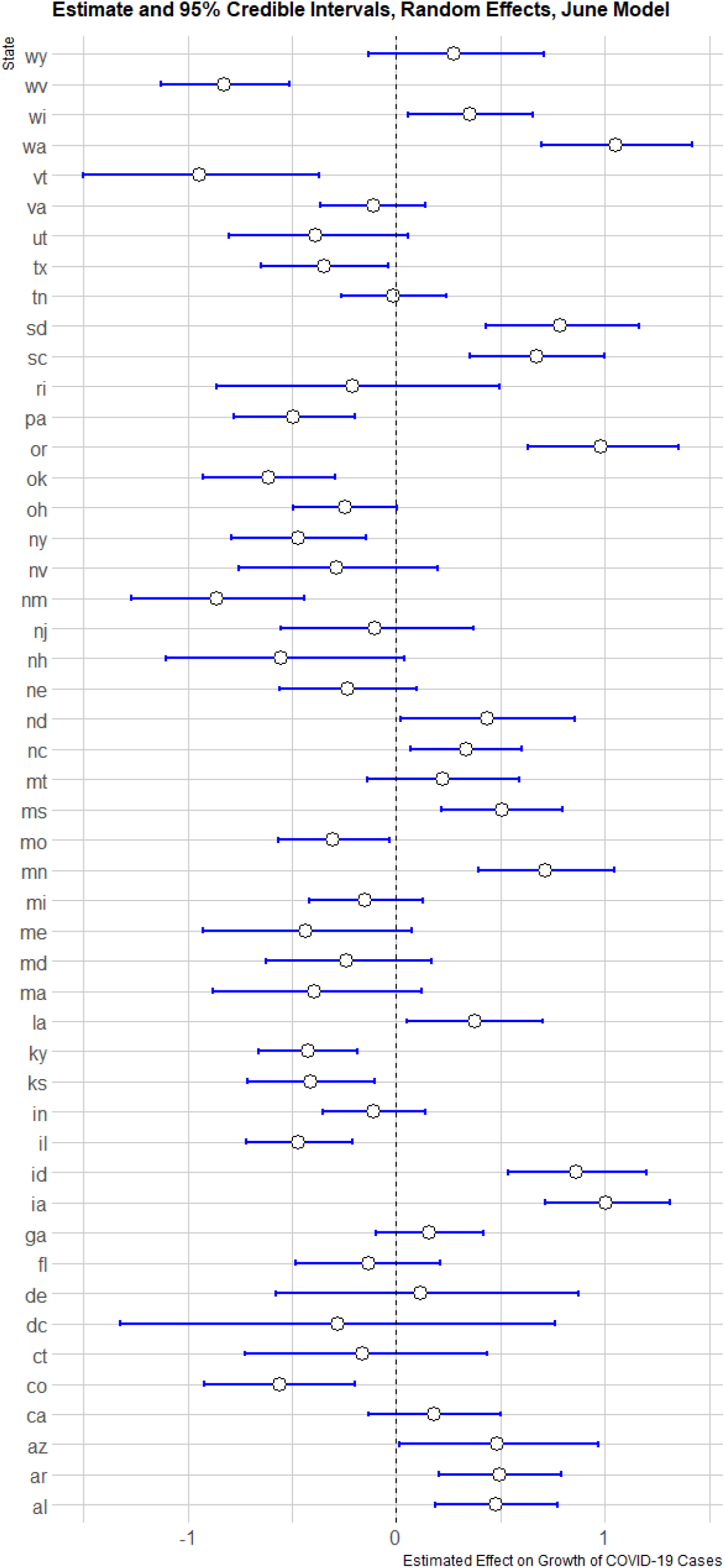
Random Effects for COVID Spread in States, late June 2020

### 3.2. Summer model

Findings from period two (late June, Table 2) present some important changes in predictors, as well as some important continuities.

**Table 1:** Predictors of SARS-COV-2 Spread, March 2020

|  | *Dependent variable:* |
| --- | --- |
|  | Rate of SARS-CoV-2 Spread |
| Average Temperature | -0.025<br>(-0.044, -0.006) |
| Density | 4.85e-04<br>(0.00039, 0.00058) |
| Density Squared | -2.03e-08<br>(-3.05e-08, -1.26e-08) |
| Density Cubed | 1.88e-13<br>(1.001e-13, 3.04e-13) |
| NYC Proper | -0.738<br>(-4.902, 4.885) |
| NYC Metro | 1.348<br>(0.805, 1.932) |
| Latitude | -0.11<br>(-0.162, -0.059) |
| Distance From U.S. Center | 0.057<br>(0.038, 0.078) |
| Relative Humidity | 0.00457<br>(-0.002, 0.011) |
| Constant | -4.536<br>(-7.303, -1.759) |
| Observations | 3,105 |
| Precision for State Random Effects | 2.801 (1.667, 4.422 ) |
| $\gamma$ | 0.002 (0.00, 0.007) |
| Effective Parameters | 53.63 |
| DIC | 16105.96 |

**Table 2:** Predictors of SARS-COV-2 Spread, June 2020

|  | *Dependent variable:* |
| --- | --- |
|  | Rate of SARS-CoV-2 Spread |
| Average Temperature | 0.019<br>(0.005,0.032) |
| Density | 2.44e-04<br>(1.69e-04, 3.23e-04) |
| Density Squared | -1.06e-08<br>(-1.63e-08, -5.87e-09) |
| Density Cubed | 1.03e-13<br>(4.92e-14, 1.67e-13) |
| NYC Proper | 0.005<br>(-2.144,2.795) |
| NYC Metro | -0.482<br>(-0.987,0.059) |
| Latitude | -0.119<br>(-0.145,-0.093) |
| Distance From U.S. Center | -0.005<br>(-0.021,0.012) |
| Relative Humidity | -0.012<br>(-0.016,-0.008) |
| Constant | -2.91<br>(-4.752,-1.067) |
| Observations | 3,108 |
| Precision for State Random Effects | 3.489 (2.155, 5.62) |
| $\gamma$ | 0.004 (0.001, 0.01) |
| Effective Parameters | 53.86 |
| DIC | 27138.31 |

The New York City Metro area was no longer significantly correlated with elevated case rates in the Summer period (−0.482, 95% CI: -0.099, 0.06). Similarly, longitude was no longer statistically significant, reflecting the more generalized spread of the virus at this point in the pandemic compared with the gradual inward spread observed during the Spring phase. Latitude showed a similar inverse correlation between the two phases (−0.119, 95% CI: -0.145, -0.093).

Population density during the Summer period remained strongly positively correlated with the spread of the virus (2.44e-04, 95% CI: 1.69e-04, 3.23e-04), and the coefficients for density-squared (−1.63e-08, 95% CI: -1.63e-08, -5.87e-09) and density-cubed (1.03e-13, 95% CI: 4.92e-14, 1.67e-13) are similar to those we observed during the earlier period. This continuity from March to June is actually quite striking and gives us increased confidence that population density is a significant factor in the spread of COVID-19.

Relative humidity (−0.012, 95% CI: -0.016, -0.008) during period 2 was negatively signed after controlling for other variables, which is consistent with early published reports [3]. The coefficient for temperature, however, shifted substantially, becoming strongly positively associated with viral spread (0.019, 95% CI: 0.005, 0.032).

Estimates of viral spread unaccounted for in our Summer model are similar to those found in the Spring model. The scale of the outbreak had grown significantly by Summer (or testing availability had improved enough to better reveal the true scale of the outbreak), so residuals are larger in absolute terms. Substantial changes in states’ observed over expected rates of spread are relatively rare. Colorado and New York transition from having greater than expected rates in the Spring to lower than expected in the Summer, while North Carolina and Oregon moved in the opposite direction.

In the Summer model, a near-majority (48 %) of counties were predicted within 10 cases, 80.5% were predicted within 50 cases, and 88% were predicted within 100 cases. When residuals are scaled by population, the rate of infection spread is estimated within a half-percentage point in 98.2% of counties, and within two-tenths of a percentage point in 92.1% of counties.

### 3.3. Sensitivity analysis

For model testing, we used the Summer model to attempt to predict viral spread for the first half of July, from July 2 to July 15. We recalculated the relative humidity data for that time period, and used the average county temperature for July. When applied to a future time period (early July), the Summer (June) model predicted 52.6% of counties within 15 cases in July, 75.8% of counties within 50 cases, and 84.9% of counties within 100 cases. Put in terms of rate, it predicted 98% of counties within a half point and 87.2% of counties within two-tents of a percent.

## 4. Discussion

Population density was the strongest predictor of viral spread in both Spring and Summer of 2020.

Consistent with the findings of earlier studies, temperature was negatively correlated with viral spread in the Spring. In the Summer, however, temperature was positively correlated with viral spread. Given that viral spread is more common in indoor spaces, this shift may reflect behavior patterns affected by the differential influence of extreme temperatures on behavior[25]. In very cold weather, such as we see in northern states in March, people choose to congregate indoors, while in very hot weather, such as we see in southern states in June, people retreat to heavily air-conditioned where close contact can facilitate viral spread; the effect of these patterns on behaviors affecting infectious and non-infectious morbidity and mortality has been documented extensively in the literature[26, 27, 28, 29, 30, 31]. Viewed in this light, the temperature variable is not causal, but rather drives behaviors that are causal.

We see some evidence of this in the residuals as well. Outliers tended to occur in counties where the popular press reported so-called “super-spreader” incidents involving indoor activities. In the Spring model, for example, Jefferson County, Louisiana, and Dougherty County, Georgia were outliers, where large outbreaks had been traced to Mardi Gras[32], and a large funeral marked by close physical contact[33].

As with the first model, the Summer model suggests a well-predicted epidemic with stochastic disease “outbreaks.” As with the first model, many (though not all) of these counties have documented causes behind their particularly large outbreaks, which are in turn largely consistent with evidence supporting indoor spread in poorly ventilated indoor spaces.

These models incorporating geographic, demographic, and environmental covariates were surprisingly accurate in their prediction of county-level rates of COVID-19 spread during two periods of 2020, despite not incorporating any variables reflecting state and regional non-pharmaceutical interventions such as “safer at home” orders, mask mandates, or business shutdowns. We would caution strongly against inferences suggesting that these interventions had no effect in predicting spread, however. First, the state-level random effects and inclusion of a zero-inflation term act, to some extent, as controls for statewide policies and testing availability. About half of the states evince significant effects suggesting some meaningful variation among states which may be related to the availability of tests and particular policy interventions. Also, very little data exist on extent to which individuals or populations conform to these orders, or the extent to which variations in rates of conformity might impact regional observations in COVID-related morbidity and mortality.

This model, in part, explains disparities in the rate of spread within the US based on environmental factors. As expected, density is related to viral spread. Temperature is more complex but consistent with the hypothesis that weather driving people indoors may facilitate spread.

We believe these results may help guide policy and future research in two ways: first, these results seem to be consistent with arguments that indoor activity is a main vector for spread. As people are less willing to hold family gatherings or to dine outdoors due to temperature extremes, the virus may transmit more rapidly. Second, we are forced to observe that viral spread is well-explained without reference to popular interventions based entirely on variables reflecting geographic, demographic, and meteorologic factors, to the exclusion of interventions aimed at reducing transmission.

Our findings are important because they assist policymakers in predicting where future viral outbreaks are likely to occur. Although the implementation of effective vaccines seems likely to slow the spread of the virus, outbreaks seem likely to continue to occur in the near future until the virus can be contained.

We also believe this serves as a warning. Winter is coming, as they say, conditions that drive people indoors can bring factors associated with increased spread perhaps giving rise to a second waves in the late fall and winter. Future research needs is warranted to assess the additional impact of targeted interventions as well as super-spreading events and behavioral variation.

## Data Availability

All data are publicly available, and references with links are provided in the manuscript.

## References

[1] E. Dong, H. Du, L. Gardner, An interactive web-based dashboard to track COVID-19 in real time, The Lancet Infectious Diseases 20 (5) (2020) 533–534. doi:10.1016/S1473-3099(20)30120-1. URL https://linkinghub.elsevier.com/retrieve/pii/S1473309920301201

[2] F. Collins, Will Warm Weather Slow Spread of Novel Coronavirus?, library Catalog: directors-blog.nih.gov (Jun. 2020). URL https://directorsblog.nih.gov/2020/06/02/will-warm-weather-slow-spread-of-novel-coronavirus

[3] J. Wang, K. Tang, K. Feng, X. Lin, W. Lv, K. Chen, F. Wang, High Temperature and High Humidity Reduce the Transmission of COVID-19, SSRN Electronic Journal (2020). doi:10.2139/ssrn.3551767. URL https://www.ssrn.com/abstract=3551767

[4] M. B. Araujo, B. Naimi, Spread of SARS-CoV-2 Coronavirus likely to be constrained by climate, preprint, Epidemiology (Mar. 2020). doi:10.1101/2020.03.12.20034728. URL http://medrxiv.org/lookup/doi/10.1101/2020.03.12.20034728

[5] R. A. Neher, R. Dyrdak, V. Druelle, E. B. Hodcroft, J. Albert, Potential impact of seasonal forcing on a SARS-CoV-2 pandemic, medRxiv (2020) 2020.02.13.20022806Publisher: Cold Spring Harbor Laboratory Press. doi:10.1101/2020.02.13.20022806. URL https://www.medrxiv.org/content/10.1101/2020.02.13.20022806v2

[6] S. Soucheray, 2020, Hospitalizations up in Arizona, Texas, Florida as US nears 3 million COVID-19 cases, CIDRAPLibrary Catalog: www.cidrap.umn.edu. URL https://www.cidrap.umn.edu/news-perspective/2020/07/hospitalizations-arizona-texas-florida-us-nears-3-million-covid-19-cases

[7] A. Ahmed, A. Kurmanaev, D. Politi, E. Londoño, Virus Gains Steam Across Latin America, The New York Times (Jun. 2020). URL https://www.nytimes.com/2020/06/23/world/americas/coronavirus-brazil-mexico-peru-chile-uruguay.html

[8] Proclamation on Declaring a National Emergency Concerning the Novel Coronavirus Disease (COVID-19) Outbreak, library Catalog: www.whitehouse.gov. URL https://www.whitehouse.gov/presidential-actions/proclamation-declaring-national-emergency-concerning-novel-coronavirus-disease-covid-19

[9] M. Holcombe, Some schools closed for coronavirus in US are not going back for the rest of the academic year, library Catalog: edition.cnn.com. URL https://www.cnn.com/2020/03/18/us/coronavirus-schools-not-going-back-year/index.html

[10] K. Zezima, J. Achenbach, T. Craig, L. H. Sun, Coronavirus is shutting down American life as states try to battle outbreak, Washington Post. URL https://www.washingtonpost.com/national/coronavirus-outbreak-shutdown-america/2020/03/13/d8589434-6550-11ea-acca-80c22bbee96f_story.html

[11] Washington Post Staff, Where states reopened and cases spiked after the U.S. shutdown, library Catalog: www.washingtonpost.com. URL https://www.washingtonpost.com/graphics/2020/national/states-reopening-coronavirus-map/

[12] E. Dong, H. Du, L. Gardner, An interactive web-based dashboard to track COVID-19 in real time, The Lancet Infectious Diseases 20 (5) (2020) 533–534. doi:10.1016/S1473-3099(20)30120-1. URL https://linkinghub.elsevier.com/retrieve/pii/S1473309920301201

[13] New York State Dept of Health, Workbook: NYS-COVID19-Tracker. URL https://covid19tracker.health.ny.gov/views/NYS-COVID19-Tracker/NYSDOHCOVID-19Tracker-Map?%3Aembed=yes&%3Atoolbar=no&%3Atabs=n

[14] US Census Bureau, American Community Survey, Tech. rep. (2019). URL https://www.census.gov/programs-surveys/acs

[15] US Census Bureau, Topologically Integrated Geographic Encoding and Referencing [TIGER] database, Tech. rep. URL https://tigerweb.geo.census.gov/tigerwebmain/TIGERwebmain.html

[16] National Centers for Environmental Information, Land-based datasets and products, Tech. rep., National Oceanic and Atmosphereic Administration. URL https://www.ncdc.noaa.gov/data-access/land-based-station-data/land-based-datasets

[17] O. A. Alduchov, R. E. Eskridge, Improved Magnus Form Approximation of Saturation Vapor Pressure, Journal of Applied Meteorology 35 (4) (1996) 601–609, publisher: American Meteorological Society. doi:10.1175/1520-0450(1996)035<0601:IMFAOS>2.0.CO;2. URL https://journals.ametsoc.org/jamc/article/35/4/601/15287/Improved-Magnus-Form-Approximation-of-Saturation

[18] P. Zhai, Y. Ding, X. Wu, J. Long, Y. Zhong, Y. Li, The epidemiology, diagnosis and treatment of COVID-19, International Journal of Antimicrobial Agents 55 (5) (2020) 105955. doi:10.1016/j.ijantimicag.2020.105955. URL https://linkinghub.elsevier.com/retrieve/pii/S0924857920301059

[19] E. K. Stokes, Coronavirus Disease 2019 Case Surveillance — United States, January 22–May 30, 2020, MMWR. Morbidity and Mortality Weekly Report 69 (2020). doi:10.15585/mmwr.mm6924e2. URL https://www.cdc.gov/mmwr/volumes/69/wr/mm6924e2.htm

[20] J. R. Fauver, M. E. Petrone, E. B. Hodcroft, K. Shioda, H. Y. Ehrlich, A. G. Watts, C. B. F. Vogels, A. F. Brito, T. Alpert, A. Muyombwe, J. Razeq, R. Downing, N. R. Cheemarla, A. L. Wyllie, C. C. Kalinich, M. Ott, J. Quick, N. J. Loman, K. M. Neugebauer, A. L. Greninger, K. R. Jerome, P. Roychoudhury, H. Xie, L. Shrestha, M.-L. Huang, V. E. Pitzer, A. Iwasaki, S. B. Omer, K. Khan, I. I. Bogoch, R. A. Martinello, E. F. Foxman, M. L. Landry, R. A. Neher, A. I. Ko, N. D. Grubaugh, Coast-to-Coast Spread of SARS-CoV-2 during the Early Epidemic in the United States, Cell 181 (5) (2020) 990–996.e5. doi: 10.1016/j.cell.2020.04.021. URL http://www.sciencedirect.com/science/article/pii/S0092867420304840

[21] H. Rue, S. Martino, N. Chopin, Approximate Bayesian inference for latent Gaussian models by using integrated nested Laplace approximations, Journal of the Royal Statistical Society: Series B (Statistical Methodology) 71 (2) (2009) 319–392. doi:10.1111/j.1467-9868.2008.00700.x. URL http://doi.wiley.com/10.1111/j.1467-9868.2008.00700.x

[22] H. Rue, A. Riebler, S. H. Sørbye, J. B. Illian, D. P. Simpson, F. K. Lindgren, Bayesian Computing with INLA: A Review, Annual Review of Statistics and Its Application 4 (1) (2017) 395–421. doi:10.1146/annurev-statistics-060116-054045. URL http://www.annualreviews.org/doi/10.1146/annurev-statistics-060116-054045

[23] J. A. Nelder, R. W. M. Wedderburn, Generalized Linear Models, Journal of the Royal Statistical Society. Series A (General) 135 (3) (1972) 370–384, publisher: [Royal Statistical Society, Wiley]. doi:10.2307/2344614. URL https://www.jstor.org/stable/2344614

[24] R Core Team, A Language and Environment for Statistical Computing (2013).

[25] R. K. Bhagat, M. S. Davies Wykes, S. B. Dalziel, P. F. Linden, Effects of ventilation on the indoor spread of covid-19, Journal of Fluid Mechanics 903 (2020) F1. doi:10.1017/jfm.2020.720.

[26] E. Cruz, S. J. D’Alessio, L. Stolzenberg, The effect of maximum daily temperature on outdoor violence, Crime & Delinquency 0 (0) (0) 0011128720926119. arXiv:https://doi.org/10.1177/0011128720926119, doi:10.1177/0011128720926119. URL https://doi.org/10.1177/0011128720926119

[27] T. M. Nelson, T. H. Nilsson, M. Johnson, interaction of temperature, illuminance and apparent time on sedentary work fatigue, Ergonomics 27 (1) (1984) 89–101, pMID: 6142820. arXiv:https://doi.org/10.1080/00140138408963466, doi:10.1080/00140138408963466. URL https://doi.org/10.1080/00140138408963466

[28] S. Wong, A. Cantoral, M. M. Téllez-Rojo, I. Pantic, E. Oken, K. Svensson, M. Dorman, I. Gutiérrez-Avila, J. Rush, N. McRae, R. O. Wright, A. A. Baccarelli, I. Kloog, A. C. Just, Associations between daily ambient temperature and sedentary time among children 4–6 years old in mexico city, PLOS ONE 15 (10) (2020) 1–21. doi:10.1371/journal.pone.0241446. URL https://doi.org/10.1371/journal.pone.0241446

[29] T. Newell, Linking outdoor air temperature and sars-cov-2 transmission in the us using a two parameter transmission model, medRxiv (2020). arXiv:https://www.medrxiv.org/content/early/2020/07/25/2020.07.20.20158238.full.pdf, doi:10.1101/2020.07.20.20158238. URL https://www.medrxiv.org/content/early/2020/07/25/2020.07.20.20158238

[30] A. I. Barreca, J. P. Shimshack, Absolute humidity, temperature, and influenza mortality: 30 years of county-level evidence from the united states, American Journal of Epidemiology 176 (2012) S114–S122. doi:10.1093/aje/kws259.

[31] D. Hruby, How an Austrian ski resort helped coronavirus spread across Europe, library Catalog: edition.cnn.com. URL https://www.cnn.com/2020/03/24/europe/austria-ski-resort-ischgl-coronavirus-intl/index.html

[32] R. A. Vargas, CDC: Mardi Gras quickened spread of coronavirus in Louisiana; canceling was never recommended, library Catalog: www.nola.com. URL https://www.nola.com/news/coronavirus/article/_dedfb5e4-7c2a-11ea-901f-6720fa25be5a.html

[33] E. Barry, Days After a Funeral in a Georgia Town, Coronavirus ‘Hit Like a Bomb’, The New York Times (Mar. 2020). URL https://www.nytimes.com/2020/03/30/us/coronavirus-funeral-albany-georgia. html

